# Comparable safety and humoral immunogenicity of delayed versus delayed fractional boosting with blood-stage malaria RH5.1/Matrix-M vaccine

**DOI:** 10.64898/2026.04.10.26348898

**Authors:** Kyra Holliday, Carolyn M. Nielsen, Thomas W. Roberts, Ellie C. Baker, Benjamin Marshall, Claire Jarman, Isaac Odongo, Jo Salkeld, Ababacar Diouf, Natalie G. Marchevsky, Rebecca Ashfield, Lloyd D. W. King, Rachel E. Cowan, Priya Lata, Fay L. Nugent, Jee-Sun Cho, Cecilia Carnot, Carole A. Long, Philip Hope, Jose Schutter, Linda Kay, Timothy Winks, Katherine Skinner, Sarah E. Silk, Simon J. Draper, Angela M. Minassian, Ruth O. Payne

## Abstract

An efficacious blood-stage malaria vaccine would serve as a highly useful public health tool alongside licensed vaccines targeting the pre-erythrocytic life cycle stage of the *Plasmodium falciparum* parasite. RH5 is the leading blood-stage malaria vaccine candidate antigen due to its highly-conserved sequence and non-redundant role in merozoite invasion of red blood cells. Following encouraging immunogenicity data in UK and Tanzanian Phase Ia/b vaccine trials, RH5-based vaccines have progressed to Phase IIb evaluation in Burkina Faso in recent years. Here, we report a Phase Ia clinical trial in malaria-naïve UK adults to assess the safety and immunogenicity of the malaria vaccine candidate RH5.1 soluble protein with Matrix-M^®^ adjuvant using two different booster dosing regimens: 10-10-10 µg versus 50-50-10 µg RH5.1, both delivered in a 0-1-6-month schedule with 50µg Matrix-M^®^ adjuvant per dose (ClinicalTrials.gov NCT06141057). A total of n=24 participants were recruited to this study, with n=23 completing all follow-up visits through to 1 year following final vaccination. The RH5.1/Matrix-M^®^ formulation was well-tolerated in this population, with injection site pain, myalgia and fatigue being the most commonly reported symptoms up to 7 days post-vaccination. There were no serious adverse events, adverse events of special interest, or suspected unexpected serious adverse reactions reported over the course of the trial. Both vaccination regimens were similarly immunogenic; no differences were observed in peak anti-RH5.1 serum IgG concentrations, *in vitro* functional anti-parasitic activity, avidity, or durability. Our findings build on other observations from clinical trials of adjuvanted RH5.1 indicating that humoral immunogenicity can be enhanced by delaying the final booster vaccination, but that there is limited impact of fractionation of the final dose. These insights can help to guide the next steps of multi-antigen, multi-stage malaria vaccine development in malaria-endemic settings.

## 1. Introduction

Malaria continues to pose a significant global health burden, causing an estimated 282 million cases and 610,000 deaths worldwide in 2024 (WHO, 2025). 75% of these deaths occurred in children under five (Villavicencio *et al*., 2024). These figures represent a plateau in previous progress since 2000. The disease is caused by the *Plasmodium* parasite and transmitted by the female *Anopheles* mosquito, with *Plasmodium falciparum* being the most prevalent species and the primary driver of human malaria morbidity and mortality.

Two recently licensed vaccines, RTS,S/AS01e (Mosquirix™) and R21/Matrix-M target the circumsporozoite protein (CSP) on the sporozoite surface to prevent invasion of hepatocytes. However, these pre-erythrocytic vaccines do not confer protection once sporozoites invade hepatocytes, leaving the host vulnerable to infection if sterile efficacy is not achieved.

A blood-stage vaccine could address this critical gap. This approach aims to protect against clinical and severe malaria while potentially allowing controlled exposure to the infection, theoretically fostering a broader and more robust adaptive immune response over time (Goodman and Draper, 2010). Such a strategy could provide children with a level of protection against disease similar to that observed in adults following repeated natural exposure. A blood-stage vaccine could be utilised independently or in combination with pre-erythrocytic or vector-stage vaccines to clear breakthrough parasitaemia and potentially induce broader immunity.

*P. falciparum* reticulocyte-binding protein homologue 5 (PfRH5) binding to basigin on the erythrocyte surface is an essential interaction that allows the merozoite to enter the red blood cell (RBC)(Crosnier *et al*., 2011). PfRH5 is part of a wider pentameric invasion complex with four other proteins including RH5-interacting protein (Ripr) and cysteine rich protein antigen (CyRPA)(Ragotte, Higgins and Draper, 2020; Scally *et al*., 2022). The process of a merozoite entering an erythrocyte is incredibly rapid, usually taking less than 30 seconds. Blocking invasion of RBCs therefore requires very high antibody titres, and previous work has demonstrated that protein-in-adjuvant formulations with highly immunogenic adjuvants such as AS01_B_ induce much higher antibody titres than viral vectored vaccines (Payne *et al*., 2017; Minassian *et al*., 2021). Unlike previous blood-stage vaccine candidate antigens such as AMA1 and MSP1, RH5 is highly conserved across different strains of *P. falciparum* making it an attractive vaccine candidate (Crosnier *et al*., 2011).

The RH5.1/Matrix-M^®^ formulation is the most advanced blood-stage vaccine candidate to date, demonstrating promising results in both Phase Ib (NCT04318002; Tanzania (Silk *et al*., 2024)) and Phase IIb (NCT05790889; Burkina Faso (Natama *et al*., 2024)) clinical trials. Further multi-antigen and/or multi-stage vaccine trials are also underway including this RH5.1/Matrix-M candidate (NCT06958198, Burkina Faso; PACTR202407803870883; Tanzania). A previous trial with RH5.1/AS01_B_ in healthy malaria-naïve UK adults strongly suggested that anti-RH5.1 serum IgG durability and B cell immunogenicity could be improved with delayed fractional (50-50-10µg at 0-1-6-months) versus monthly (2-2-2 or 10-10-10 or 50-50-50µg at 0-1-2-months) booster RH5.1 protein dosing regimens that used a fixed 0.5mL dose of AS01_B_ (Minassian *et al*., 2021; Nielsen *et al*., 2021). Delayed booster dosing has therefore subsequently been explored with the RH5.1/Matrix-M candidate in the above Phase Ib and Phase IIb trials in malaria endemic regions and, again, has shown a positive impact on

IgG and B-cell immunogenicity (Barrett, Pipini, *et al*., 2024; Natama *et al*., 2024; Silk *et al*., 2024)). However, while both these RH5.1/Matrix-M trials broadly support the use of delayed booster dosing (i.e. 0-1-6 month or 0-1-5 month versus 0-1-2 month), there are no data comparing delayed versus delayed fractional regimens (10-10-10µg vs 50-50-10µg RH5.1, respectively) on a 0-1-6-month vaccination schedule in healthy adults. In this current trial (NCT06141057), we address this data gap and provide a direct comparison of a delayed fractional booster (50-50-10µg; DFx) to a standard dose delayed booster regimen (10-10-10µg; 10D) on a 0-1-6-month schedule for the RH5.1/Matrix-M vaccine candidate in healthy adults.

For administration, 50μg Matrix-M^®^ adjuvant, a potent saponin-based adjuvant from Novavax AB (Uppsala, Sweden), was mixed with the RH5.1 in the clinical research facility, prior to administration. This adjuvant has a well-established safety profile, and is used in licensed COVID-19 and malaria vaccines and experimentally in other vaccines (Heath *et al*., 2021; Datoo *et al*., 2024; Shinde *et al*., 2022; Stertman *et al*., 2023).

The primary objective was to establish the safety of the RH5.1/Matrix-M vaccine in healthy UK adults with these two booster schedules. The secondary objectives were to: a) assess humoral immunogenicity of RH5.1/Matrix-M in each booster dosing regimen; b) compare magnitude, functionality, and longevity of anti-RH5.1 serum IgG between dosing regimens; and c) compare differences in innate immune responses following the first and third vaccinations and correlate these with adverse event data and adaptive immune responses. Here, we report on the core safety and anti-RH5.1 serum IgG endpoints; innate analyses will be the focus of a separate analysis (Holliday *et al*., *manuscript in preparation*).

## 2. Materials and Methods

### 2.1 RH5.1/Matrix-M vaccine

The design, production and preclinical testing of the RH5.1 protein vaccine have been previously reported in detail (Jin *et al*., 2018). Briefly, RH5.1 is a soluble protein immunogen based on the full-length PfRH5. The protein is produced by secretion from *Drosophila melanogaster* Schneider-2 cells provided by ExpreS2ion Biotechnologies (Denmark). RH5.1 was developed at the University of Oxford and an initial batch, produced to Good Manufacturing Practice (GMP) by the Clinical Biomanufacturing Facility in Oxford, was used in previously reported clinical trials VAC063 (Minassian *et al*., 2021) and VAC080 (Silk *et al*., 2024). Using the same established manufacturing processes, a second clinical batch of RH5.1 was filled in 2021 under GMP by a Contract Manufacturing Organisation based in the UK. This new batch of vaccine was used in the BIO-002 clinical trial reported here.

### 2.2 Trial design

This study, BIO-002, was a single-blind, randomised, Phase Ia clinical trial evaluating the safety and immunogenicity of the protein-in-adjuvant vaccine candidate RH5.1/Matrix-M in malaria-naïve UK adults. Participants were blinded to the dose received and randomised 1:1, stratified only by biological sex, to two different dosing schedules: three 10µg doses RH5.1 (’10D’) or two 50µg doses followed by one 10µg dose (’DFx’). All were adjuvanted with 50µg Matrix-M and administered by intramuscular injection at 0, 1, and 6 months. A custom REDCap database was used to generate the randomisation after eligibility was confirmed. The study was conducted at the NIHR Clinical Research Facility (CRF) in Sheffield Teaching Hospitals in collaboration with the University of Sheffield (Sheffield, UK) and University of Oxford (Oxford, UK). The study was sponsored by the University of Oxford. The trial was registered with ClinicalTrials.gov (NCT06141057) and ISRCTN95289709. The primary endpoint of the study was the safety and reactogenicity of RH5.1 and Matrix-M, with secondary immunogenicity outcomes (see **Supplementary Methods** for more detail).

Healthy, malaria-naïve male and non-pregnant female volunteers aged 18-50 were invited to participate in the study. Volunteers were consented, recruited and vaccinated at the CRF at the Royal Hallamshire Hospital, Sheffield. Twenty-four volunteers were recruited. A full list of inclusion and exclusion criteria is available in Error! Reference source not found..

The study received approval from London-Chelsea Research Ethics Committee (REC reference 23/LO/0058), the Medicines & Healthcare products Regulatory Agency (MHRA) and Health Research Authority (HRA) via the Integrated Research Application System (IRAS) (IRAS Project ID 1005754). Volunteers signed written consent forms, and consent was verified before each vaccination. The trial was conducted according to the principles of the current revision of the Declaration of Helsinki 2013 and in full conformity with the ICH guidelines for Good Clinical Practice (GCP).

### 2.3 Safety analysis

Following each vaccination volunteers completed an electronic diary card daily for seven days via REDCap (day of vaccination and for six days post vaccination), recording solicited adverse events (AEs), which included temperature, local swelling or redness (mm), pain, itching or warmth and generalised symptoms of feverishness, myalgia, arthralgia, headache, fatigue, nausea and malaise. AEs occurring after seven days or not included as solicited AEs were recorded for 28 days following each vaccination as ‘unsolicited AEs’. All AEs were graded for severity by the volunteers and causality relating to vaccination was assessed by investigators. Physical observations (blood pressure, heart rate and temperature) were recorded at all study visits. Blood samples for safety (full blood count, liver function, urea and electrolytes) were performed at screening, Day 0 (D0), D1, D2, D7, D28, D29, D35, D182, D183, D184, D189 and D210 for all groups. Serious Adverse Events (SAEs) and Adverse Events of Special Interest (AESIs) were reviewed at all study visits. All AEs considered to be possibly, probably or definitely related to vaccination are reported. Further details on the grading and causality of AEs and the defined AESIs are in the Supplementary Methods (Tables S3, S4 and S5).

### 2.4. Total IgG and isotype ELISAs

Standardised ELISAs were used to quantify the magnitude of anti-RH5.1 serum IgG pre- and post-vaccination with RH5.1/Matrix-M at all study visits other than screening. Anti-RH5.1 total IgG ELISAs were performed against full-length RH5 protein (RH5.1) using standardised methodology as previously described (Minassian *et al*., 2021; Silk *et al*., 2023, 2024). The reciprocal of the test sample dilution giving an OD_405_ of 1.0 in the standardised assay was used to assign an ELISA unit value of the standard. A standard curve and Gen5 ELISA software v3.04 (BioTek, UK) were used to convert the OD_405_ of individual test samples into AU. AU were converted into μg/mL following generation of a conversion factor by calibration-free concentration analysis as reported previously (Payne *et al*., 2017).

Standardised ELISAs were similarly performed to quantify the magnitude of anti-RH5.1 IgG1, IgG3, IgG4, IgA and IgM responses (Payne *et al*., 2017; Minassian *et al*., 2021; Barrett, Silk, *et al*., 2024). In brief, Nunc MaxiSorp™ flat-bottom ELISA plates (44-2404-21, Invitrogen) were coated overnight with 5µg/mL RH5.1 protein in PBS. Plates were washed with washing buffer composed of PBS containing 0.05% TWEEN® 20 (P1379, Sigma-Aldrich) and blocked with 100µL Starting Block™ T20 (37538, ThermoFisher Scientific). After removing the blocking buffer, standard curve and internal controls were diluted in blocking buffer using a pool of high-titre vaccinee plasma from a previous RH5.1/AS01_B_ clinical trial (NCT02927145), and 50µL each dilution was added to the plate in duplicate. Test samples were diluted in blocking buffer to a minimum dilution of 1:50 and 50µL was added in triplicate. Plates were incubated for 2h at 37°C and washed in washing buffer. An alkaline phosphatase-conjugated secondary antibody was diluted at the manufacturer’s recommended minimum dilution for ELISA in blocking buffer. The antibody used was dependent on the isotype or subclass being assayed and were as follows: IgG1 Hinge-AP (9052-04, Southern Biotech); IgG3 Hinge-AP (9210-04, Southern Biotech); IgG4 Fc-AP (9200-04, Southern Biotech); IgA-AP (2050-04, Southern Biotech); and IgM-AP (2020-04, Southern Biotech). Then, 50µL of the secondary antibody dilution was added to each well of the plate and incubated for 1h at 37°C. Plates were developed using PNPP alkaline phosphatase substrate (N2765, Sigma-Aldrich) for 1-4h at 37°C. Optical density at 405 nm was measured using an ELx808 absorbance reader (BioTek) until the internal control reached an OD_405_ of 1. The reciprocal of the internal control dilution giving an OD_405_ of 1 was used to assign an AU value of the standard. Gen5 ELISA software v3.04 (BioTek) was used to convert the OD_405_ of test samples into AU values by interpolating from the linear range of the standard curve fitted to a four-parameter logistic model. Any samples with an OD_405_ below the linear range of the standard curve at the minimum dilution tested were assigned a minimum AU value according to the lower limit of quantification of the assay.

### 2.5 Avidity ELISA

Anti-RH5.1 total IgG antibody avidity was assessed by sodium thiocyanate (NaSCN)-displacement ELISA using previously described methodology (Payne *et al*., 2017; Minassian *et al*., 2021; Silk *et al*., 2023). The concentration of NaSCN required to reduce the OD_405_ to 50% of that without NaSCN was used to report a measure of avidity (IC_50_).

### 2.6 Growth inhibition activity (GI) assay

Standardised assays were performed by the GIA Reference Center, NIH, USA, using previously described methodology (Malkin *et al*., 2005). Operators performing the GIA assays were blinded to participant group allocation. Each sample was tested in three independent replication assays using three different batches of red blood cells (RBC), and the median of these three results was used to generate the final dataset as per previously reported RH5 vaccine trials and methodology (Miura *et al*., 2009; Bustamante *et al*., 2013; Osier *et al*., 2014) For each assay, protein G purified IgG samples were incubated with RBC infected with synchronised *P. falciparum* 3D7 clone parasites in a final volume of 40μL for 40 h at 37 °C, and the final parasitaemia in each well was quantified by biochemical determination of parasite lactate dehydrogenase. All purified IgG samples were tested initially at 10 mg/mL final test well concentration. For samples with greater than 20% GIA at 10 mg/mL, a dilution series was tested and used to determine the concentration that gave 50% GIA (EC_50_).

### 2.7 Statistical Analyses

The sample size of 12 participants per group was determined pragmatically, without formal power calculations. All analyses for the primary safety outcome were descriptive, with no hypothesis testing performed.

All safety events were reported by group and dose. Solicited and laboratory AEs were summarised by frequencies and proportions of participants experiencing events, by severity; unsolicited AEs deemed possibly, probably, or definitely related to the study vaccine were summarised by frequency and maximum severity of events.

For immunology analyses, data were analysed using GraphPad Prism version 10 for Windows (GraphPad Software Inc). Limited statistical comparisons were conducted between groups given the small sample size and consequent lack of statistical power. For all formal hypothesis tests, the type I error rate was set at 0.05 and Mann-Whitney *U* testing was used. Except when there were three groups to compare when including a historical cohort, where Kruskal-Wallis with Dunn’s test of multiple comparisons was applied. P values were adjusted to account for multiple comparisons where appropriate, as indicated in figure legends. We note that non-significant findings cannot be interpreted to mean there is no difference between groups, and significant findings may not be replicable.

Solicited AE data were visualised using R version 4.4.2 (2024-11-01) in RStudio.

## 3. Results

### 3.1 Participant demographics and follow-up

From 25th September 2023 to 2nd February 2024, 38 UK adult volunteers were screened in total and 24 healthy adult volunteers were enrolled and randomised into two groups of 12. The participant demographics are summarised in **Table 1**. There were more female (66%) than male (33%) participants in the study, with the same proportions in each group. The median age of participants across the study was 37.5 years (IQR 28-44), and the majority of participants were from a white ethnic background.

**Table 1.** Baseline demographics of study groups.

|  | 10D group (n=12) | DFx group (n=12) | Overall (n=24) |
| --- | --- | --- | --- |
| Median age (years) | 40 (27.5-44) | 36 (31-44.5) | 37.5 (28-44) |
| Range | 24-48 | 24-49 | 24-49 |
| **Sex (n,%)** |  |  |  |
| Male | 4 (33%) | 4 (33%) | 8 (33%) |
| Female | 8 (66%) | 8 (66%) | 16 (66%) |
| **Ethnicity (n,%)** |  |  |  |
| White British | 10 (83.3%) | 11 (91.7%) | 21 (87.5%) |
| Any other White | 1 (8.3%) | 1 (8.3%) | 2 (8.3%) |
| Pakistani | 1 (8.3%) | 0 (0%) | 1 (4.2%) |

The 10D group received three 10µg RH5.1 vaccinations at 0, 1 and 6 months. The DFx group received two higher dose vaccinations of 50µg RH5.1 at 0 and 1 month, followed by a 10µg dose vaccination at 6 months. All RH5.1 vaccinations were administered with 50µg Matrix-M^®^ adjuvant. Vaccinations began on 23rd October 2023 and all follow up visits were completed by 16th July 2025. One volunteer withdrew due to personal reasons after the Day 35 visit and was not replaced; all other participants completed the vaccinations and follow up as planned (**Figure 1**).

**Figure 1.**
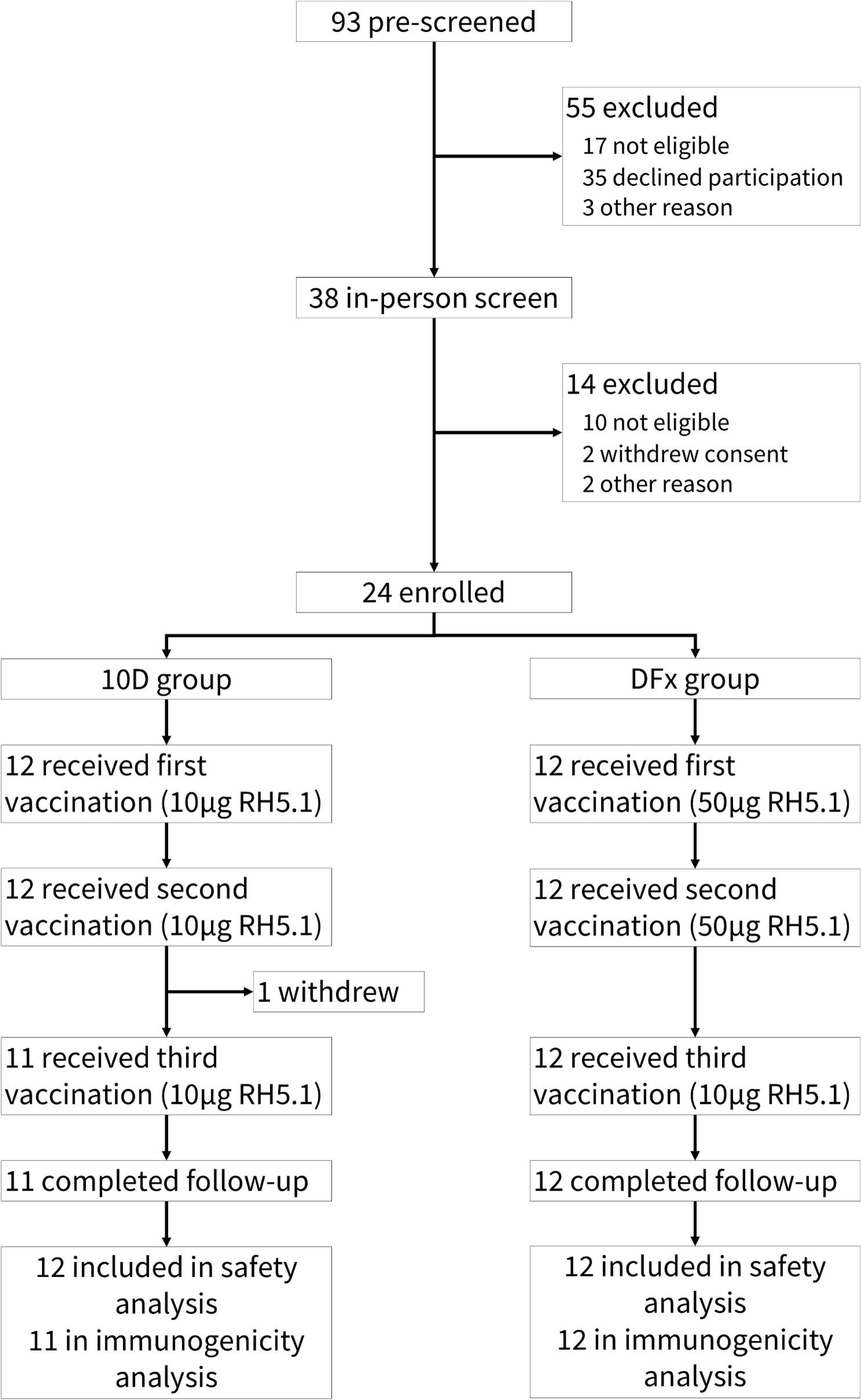
BIO-002 CONSORT diagram. All RH5.1 vaccinations were administered with 50µg Matrix-M® adjuvant. One participant in the 10D group withdrew after the second study vaccination. This was due to life events and not due to adverse events or reactions. All participants who received at least one dose were included in the safety analysis. Pre-screening was performed via an online questionnaire.

### 3.2 Solicited adverse events (AEs)

Pain was the most commonly reported local AE, occurring after 76% (54 of 71) of all doses administered. The majority of these (63%, 34 of 54) were reported after the second and third doses. All local solicited AEs were mild or moderate in severity (**Figure 2**).

**Figure 2.**
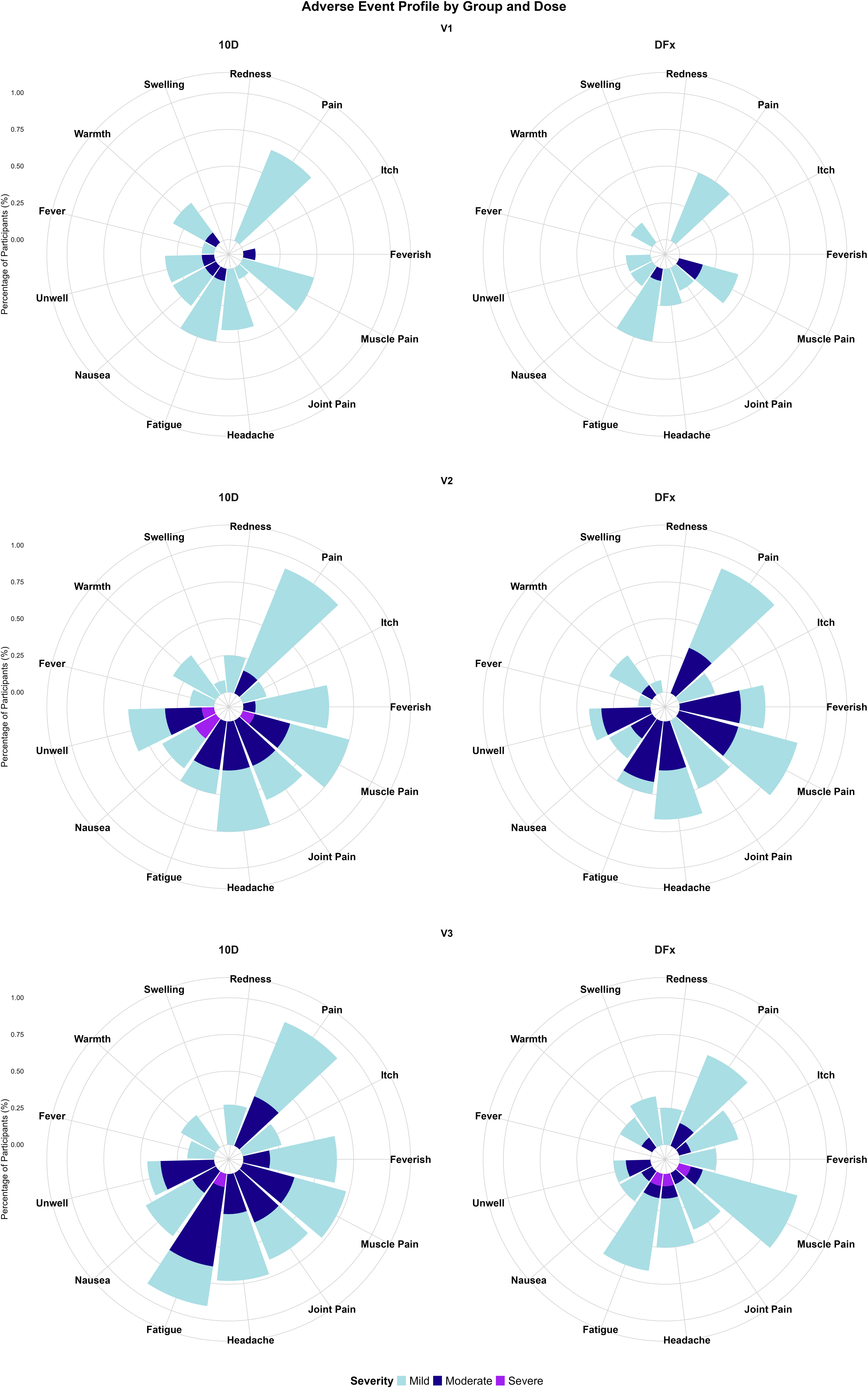
Maximum solicited AE grades after each vaccine visit shown by group. The maximum severity reported for each symptom as a percentage of that group (10D or DFx; running horizontally) and vaccination visit (V1, V2 or V3; running vertically). These were self-reported via a daily online webform on REDCap. There were no missing entries.

Systemic solicited AEs were again more prominent after the second and third doses, indicating increased reactogenicity with subsequent administrations. Most symptoms were mild or moderate in severity; with myalgia and fatigue being the most common; reported after 68% (48 of 71) and 59% (42 of 71) of all doses, respectively.

Whilst most systemic solicited AEs were mild to moderate, four participants noted five instances of severe solicited AEs, all of which occurred after the second and third doses (**Figure 2**). After the second dose in the 10D group, one participant reported severe nausea, myalgia and malaise and another severe nausea associated with a vasovagal faint; no-one in the DFx group reported severe solicited AEs. After the third dose, one participant in the 10D group reported severe fatigue. In the DFx group, one participant reported severe fatigue and myalgia and another severe headache five days post-vaccination. Severe solicited AEs resolved within 24 hours for 3 of the 4 participants and within 48 hours for the other.

Overall, subsequent doses appeared more reactogenic than the first. All solicited AEs resolved spontaneously; most within 72 hours. The only interventions used were self-administered over-the-counter analgesia (such as paracetamol, ibuprofen or co-codamol) which were used by 54% (13 of 24) of participants during the 7-day post-vaccination reporting period.

### 3.3 Unsolicited adverse events (AEs) and laboratory AEs

The RH5.1/Matrix-M vaccine candidate demonstrated an acceptable safety profile in this cohort of healthy UK adults. There were no SAEs, AESIs, or suspected unexpected serious adverse reactions (SUSARs) reported up to one year after the final vaccination.

The most frequently reported unsolicited adverse AEs were injection site reactions occurring approximately one-week post-vaccination, after Day 6 when the solicited reporting was complete. In total, five participants reported these delayed local reactions. All had itching, which was sometimes associated with erythema, warmth or swelling, whilst one was a systemic reaction presenting as generalised, all-over itching that lasted for two days (Days 7-9). A summary of the unsolicited events reported throughout the trial is available in **Table S7**. One participant also reported an intermittent rash on the trunk over a period of 227 days, starting after the first vaccine. However, this was not reported until after they had received all three doses, so the exact date of onset is unclear. This was deemed possibly related to the vaccination. This and all other events resolved spontaneously.

The most frequent laboratory abnormality was lymphocytopenia occurring in 67% (16/24) participants on samples the day after vaccine visits (D1, D29 and D183) (**Table 2**). Of those in the 10D group, more were mild in graded severity than in the DFx group where most were moderate in severity. These changes all recovered within 48 hours. One participant had a grade 3 lymphocytopenia on D29 and was brought back for a repeat sample on D32, confirming resolution.

**Table 2.** Summary of laboratory AEs. All participants with a grade 1 or higher laboratory abnormalities are reported. N=12 in each group. 10D=delayed third dose regimen; DFx=delayed-fractional third dose regimen. The defined ranges for AE grading are available in the Supplementary Methods.

| Laboratory abnormality | 10D |  |  |  | DFx |  |  |  |
| --- | --- | --- | --- | --- | --- | --- | --- | --- |
|  | No. of participants | Grade |  |  | No. of participants | Grade |  |  |
|  |  | 1 | 2 | 3 |  | 1 | 2 | 3 |
| **Lymphocytopenia** | 9 (75%) | 5 (42%) | 3 (25%) | 1 (8%) | 7 (58%) | 1 (8%) | 4 (33%) | 2 (17%) |
| **Elevated alanine Aminotransaminase** | 2 (17%) | 2 (17%) | 0 | 0 | 3 (25%) | 2 (17%) | 1 (8%) | 0 |
| **Leukopenia** | 2 (17%) | 2 (17%) | 0 | 0 | 0 | 0 | 0 | 0 |
| **Leukocytosis** | 3 (25%) | 3 (25%) | 0 | 0 | 0 | 0 | 0 | 0 |
| **Neutropenia** | 3 (25%) | 3 (25%) | 0 | 0 | 3 (25%) | 3 (25%) | 0 | 0 |
| **Anaemia** | 1 (8%) | 0 | 1 (8%) | 0 | 1 (8%) | 1 (8%) | 0 | 0 |
| **Elevated creatinine** | 1 (8%) | 1 (8%) | 0 | 0 | 2 (17%) | 2 (17%) | 0 | 0 |
| **Thrombocytopenia** | 1 (8%) | 1 (8%) | 0 | 0 | 0 | 0 | 0 | 0 |
| **Eosinophilia** | 1 (8%) | 1 (8%) | 0 | 0 | 1 (8%) | 1 (8%) | 0 | 0 |
| **Elevated bilirubin** | 1 (8%) | 1 (8%) | 0 | 0 | 0 | 0 | 0 | 0 |
| **Haemoglobin change** | 11 (92%) | 6 (50%) | 3 (25%) | 2 (17%) | 7 (58%) | 6 (50%) | 0 | 1 (8%) |

Of the five participants with elevated alanine aminotransferase (ALT), see **Table 2**, two were deemed potentially clinically significant. Neither was felt to be related to the study vaccine. This was due to the elevation being present at Day 28, prior to their second vaccination. Both had suffered from different viral illnesses (one gastroenteritis, the other upper respiratory tract infection) in the period between Day 14 and Day 28; this was over the winter period 2023/2024 and these illnesses were felt to be seasonal viruses. Both infections were self-limiting, and participants had physically recovered by the Day 28 vaccination visit, but a biochemical imbalance was still evident.

Overall, 75% (18/24) of participants had a change in haemoglobin during the trial (mainly reductions, although increases were also recorded). The majority of these changes were mild (67%; 12/18) with three moderate and two severe in the 10D group and one severe in the DFx group. The haemoglobin changes mostly occurred during the intensive period of follow up after each vaccine where up to 165ml (or around half a unit) of blood was taken over 3 days. Therefore, they are likely all related to the volume of blood being taken during the trial. Haemoglobin also has variability depending on the hydration level of the participant. Most of these participants stayed within the normal range for haemoglobin. No participants dropped into a moderate or severe absolute haemoglobin level. Those that dropped into a mild severity haemoglobin level already had low-normal haemoglobin to start. Only one of these was deemed to be clinically significant and referred to the general practitioner for follow up. This participant had been diagnosed with Type 2 Diabetes Mellitus between the second and third vaccine and started metformin which can also be associated with an early risk of anaemia (Donnelly *et al*., 2020). The new diagnosis of Type 2 Diabetes Mellitus was felt to be unrelated to vaccination. This was the only laboratory abnormality that had not resolved before the end of the trial.

### 3.4 Comparable anti-RH5.1 total IgG responses in delayed and delayed fractional booster dosing regimens with RH5.1/Matrix-M

Anti-RH5.1 serum IgG responses were detected in both 10D and DFx groups following vaccination with RH5.1/Matrix-M (**Figure 3A-B**). Responses peaked at Day 196 (14 days post-vaccination 3; V3+14) in both groups (10D median = 153 µg/mL [range 38-280 µg/mL]; DFx median = 72 µg/mL [range 18-287 µg/mL]) and declined approximately 15-fold by Day 547 (V3+1yr; 10D median = 11 µg/mL [range 4-60 µg/mL]; DFx median = 5 µg/mL [range 1-29 µg/mL]). While anti-RH5.1 IgG concentrations trended higher in the delayed boosting 10D participants at most time points as compared to delayed fractional DFx participants, these differences were not statistically significant at the key time points tested by Mann-Whitney *U* (Day 0, Day 196, Day 547).

**Figure 3.**
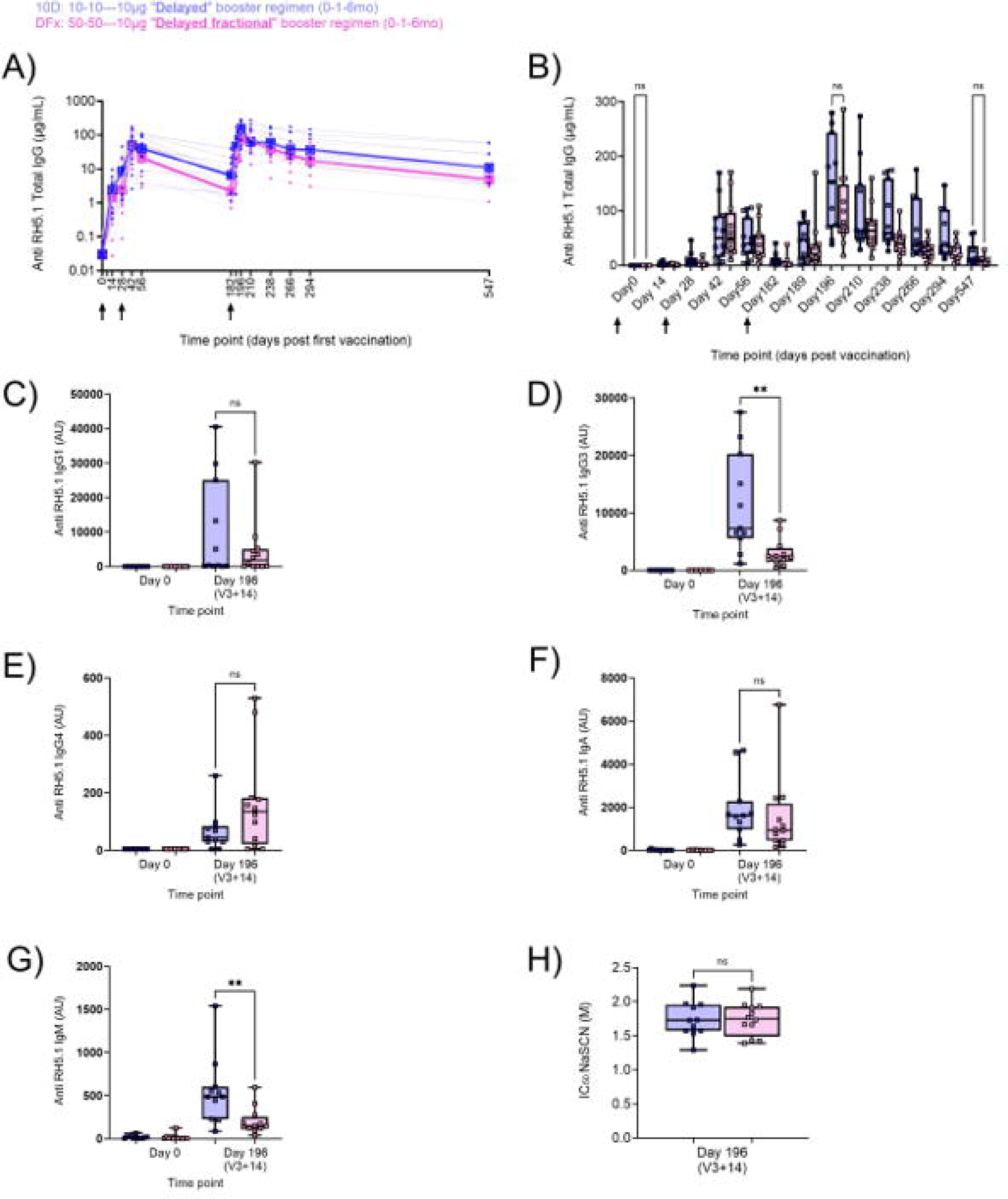
Anti-RH5.1 serum antibody responses to delayed or delayed fractional booster dosing with RH5.1/ Matrix-M®. Full anti-RH5.1 serum IgG kinetics from pre-vaccination baseline (Day 0) to 1-year post-third vaccination (Day 547) were measured by ELISA (**A-B**). A Mann-Whitney *U* test was performed to compare serum IgG concentrations between 10D and DFx participants at three key time points only: Day 0, Day 196, and Day 547 (**B**). Standardised ELISAs were similarly performed to compare anti-RH5.1 serum IgG1 (**C**), IgG3 (**D**), IgG4 (**E**), IgA (**F**) and IgM (**G**) responses at peak time point (Day 196; V3+14) between 10D and DFx participants, with analysis by Mann-Whitney *U* test. A NaSCN displacement assay was used to compare avidity of anti-RH5.1 IgG between 10D and DFx participants, with analysis by Mann-Whitney *U* test (**H**). In all graphs, Group 1 (10D) data are shown in blue and Group 2 (DFx) in pink. Each point represents a single sample with the exception of (**A**) where thick lines and large points represent group medians. In other graphs, bars and error bars represent median and interquartile ranges, respectively. In (**A-B**), arrows indicate days of vaccination (Day 0, Day 28, Day 182). p values are annotated on graphs; p < 0.05 was considered significant. ns = non-significant. ** p < 0.01. V3 = vaccination 3.

In contrast, we did observe indications of differences within RH5.1-specific antibody isotypes and subclasses between 10D and DFx participants when assayed at the peak time point (Day 196; V3+14; **Figure 3C-G**). Specifically, significantly higher anti-RH5.1 IgG3 and IgM concentrations were observed in 10D delayed boosting participants as compared to DFx delayed fractional participants by Mann-Whitney *U* test (**Figure 3D,G**).

In terms of the quality of the total IgG response, anti-RH5.1 IgG avidity was measured with a sodium thiocyanate displacement assay. Here, no significant differences were observed between 10D and DFx participants (**Figure 3H**) with median IC_50_ values observed in both groups (10D median = 1.7 M [range 1.3-2.2 M]; DFx median = 1.7 M [range 1.4-2.2 M]) similar to what we previously observed following delayed fractional dosing with RH5.1/AS01_B_ in malaria-naïve UK adults (Minassian *et al*., 2021).

While small sample sizes limit conclusive comparisons between Phase I trials, we were also interested to contextualise the anti-RH5.1 IgG immunogenicity and GIA data from the 10D and DFx RH5.1/Matrix-M vaccinees with previous reports from historical RH5.1/AS01_B_ UK adult vaccinees (Minassian *et al*., 2021). Using a Kruskal-Wallis test to compare serum anti-RH5.1 IgG concentrations between vaccine groups at baseline, peak, and “late” time-point, we observed that peak anti-RH5.1 IgG responses are not significantly different between DFx dosing with RH5.1/Matrix-M and RH5.1/AS01_B_ (Group 3 in NCT02927145), but the latter group is significantly higher at the late time point follow-up (Group 3 NCT02927145 median = 50 µg/mL [range 29-140 µg/mL]) despite the late time-point in this trial being sampled at Day 822 rather than Day 547 using Dunns multiple comparisons (**Supplemental Figure 1A**).

### 3.5 Blood-stage malaria parasite growth inhibition activity (GIA) of post-vaccination IgG with delayed and delayed fractional RH5.1/Matrix-M

To compare *in vitro* functional anti-parasitic activity of the vaccine-induced anti-RH5.1 IgG response, blinded serum samples were run at the GIA Reference Center GIA Reference Centre (National Institute of Allergy and Infectious Diseases, National Institutes of Health). Purified total IgG was normalised to a starting concentration of 10 mg/mL and tested against *P. falciparum* (3D7 clone) parasites using the standardised single-cycle GIA assay. All baseline pre-vaccination samples (Day 0) showed negligible GIA while peak post-vaccination samples (Day 196; V3+14) showed a substantial increase in activity (**Figure 4A-B**). All total IgG samples with greater than 20% GIA at 10 mg/mL were subsequently titrated using a 2-fold dilution series in the assay (**Figure 4 A-B**); this included 10/11 10D participants’ and 11/12 DFx participants’ Day 196 IgG samples. The median percentage GIA trended higher in the 10D participant post-vaccination samples at all concentrations of total IgG tested. To limit the multiplicity of testing, we only performed a formal statistical test on the 2.5 mg/mL total IgG concentration – as most commonly reported in previous RH5 vaccine clinical trials – and here we observed no significant difference between 10D and DFx by Mann-Whitney *U* (10D median = 27.7% [range -2.4-42.1%]; DFx median = 20.9% [range 4.9-44.7%]; **Figure 4B**).

**Figure 4.**
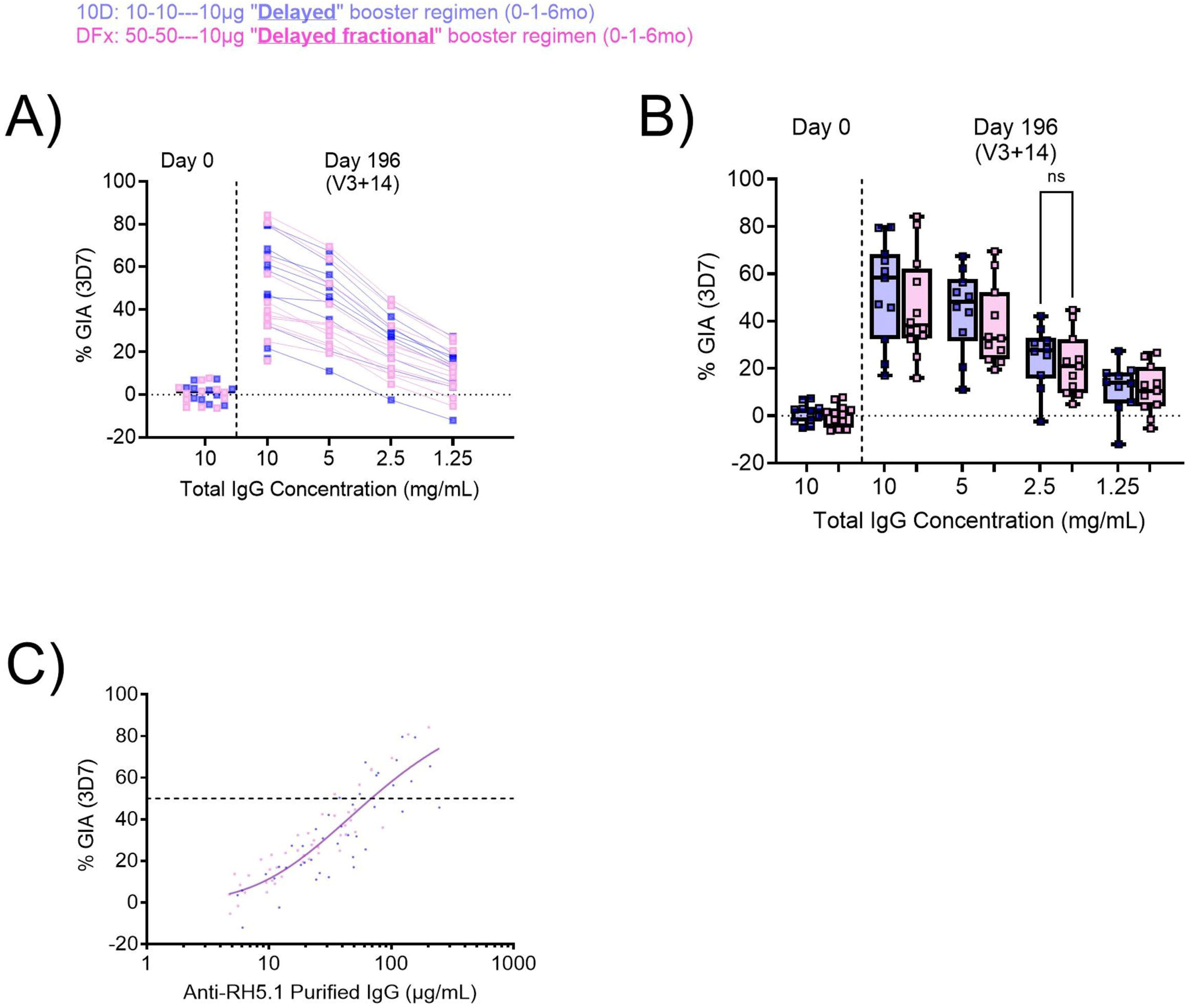
Functional parasite growth inhibition activity (GIA) induced by delayed or delayed fractional booster dosing with RH5.1/ Matrix-M^®^. *In vitro* GIA was analysed with total IgG purified from pre-vaccination baseline (Day 0) or peak post-vaccination (Day 196; V3+14) serum. All samples were screened at 10 mg/mL total IgG, followed by a titration to 5 mg/mL, 2.5 mg/mL and 1.25 mg/mL for samples with > 20% GIA at the initial 10 mg/mL screen (**A-B**). GIA activity at 2.5 mg/mL was compared between 10D and DFx participants by Mann-Whitney *U* test (**B**). The relationship between GIA data from the dilution series and the anti-RH5.1 IgG concentration as measured by ELISA in the total purified IgG samples was used to determine “quality” of the anti-RH5.1 IgG response (**C**). Non-linear five-parameter regression curves (constrained to >0% and <100 % GIA) are shown for both groups. Concentration of anti-RH5.1-specific polyclonal IgG that gives 50% GIA (EC_50_; dashed line represents 50% GIA) was calculated using data points pooled across all groups (r^2^=0.8; n=23). In all graphs, Group 1 (10D) data are shown in blue and Group 2 (DFx) in pink. Each point represents a single sample. Bars and error bars represent median and interquartile ranges, respectively (**B**). p < 0.05 was considered significant. ns = non-significant. V3 = vaccination 3.

We next assessed the concentration of RH5.1-specific IgG required to give 50% GIA (EC_50_), which is an informative read-out to allow comparison of the “quality” of the antigen-specific IgG response between clinical trials. Analysis across both groups combined showed a GIA EC_50_ of 69 µg/mL (95% confidence interval 58-80; **Figure 4C**).

Finally, as with the serology analyses above, we sought to contextualise this 2.5 mg/mL GIA data with previously reported data from RH5.1/AS01_B_ delayed fractional boosting UK adult vaccinees (DFx NCT02927145). Here, again by Mann-Whitney *U*, we observed higher GIA in these RH5.1/AS01_B_ vaccinees at the matched Day 196 (V3+14) time point (DFx NCT02927145 median GIA = 31.2% [range 22.5-62.3%]) as compared to DFx RH5.1/Matrix-M participants (**Supplemental Figure 1B**).

## 4. Discussion

This study reports the first head-to-head comparison of 10-10-10µg (delayed) and 50-50-10µg (delayed fractional) booster dosing of RH5.1/Matrix-M vaccination on a 0-1-6-month schedule in healthy adults. The results confirm that both regimens have a satisfactory safety profile and are well-tolerated in a malaria-naïve UK adult population.

Overall, the reactogenicity profile observed was acceptable and consistent with the previous RH5.1/AS01_B_ trial in UK adults (NCT02927145)(Minassian *et al*., 2021). In contrast, we did observe minor differences in the symptom profile in comparison to RH5.1/Matrix-M Tanzanian adult data (NCT04318002)(Silk *et al*., 2024). Although pain was the most common feature in both cohorts (35% Tanzanian adults; 76% UK adults); systemic symptoms were more common in this UK cohort (68% and 59% reported myalgia and fatigue, respectively in the UK cohort, compared to only 3% and 6% in Tanzania). (Silk *et al*., 2024). The proportionally higher reporting of all solicited symptoms in the study here aligns with previous similar observations when comparing malaria-naïve (UK) cohorts to malaria-endemic populations (Silk *et al*., 2023; Venkatraman *et al*., 2025). This may be due to reporting bias (Thriemer *et al*., 2023) or pre-exposure to malaria antigens causing semi-immunity that actually dampens the inflammatory response (Pohl and Cockburn, 2022). Either explanation would be of interest to understand further but is not considered by the investigators to be a cause for concern with respect to vaccine safety.

The pattern of injection site reactions beginning a week post-vaccination observed in the BIO-002 study here suggests a delayed-type hypersensitivity reaction (Type IV). These are usually T cell-mediated (Stone Jr *et al*., 2019). Importantly, all these reactions were self-limiting, resolving spontaneously with the majority lasting no more than nine days; these injection site reactions therefore do not pose a long-term safety concern. Indeed, the absence of any SAEs or AESIs up to one year post-final vaccination provides further support for the safety of this vaccine candidate, building on the previously published safety and immunogenicity data from 12 adults and 347 children across Phase Ib and IIb trials in malaria-endemic settings (Natama *et al*., 2024; Silk *et al*., 2024).

Another key finding of this study is the comparable immunogenicity of the two dosing strategies, specifically the magnitude of the peak anti-RH5.1 IgG serum concentration, as well as longevity, avidity and functional anti-parasitic activity of this anti-RH5.1 IgG. Our data suggest that previously reported improvements in adult vaccinee RH5.1 immunogenicity between monthly (0-1-2-month) and delayed fractional (0-1-6-month) booster dosing regimens (Minassian *et al*., 2021; Nielsen *et al*., 2023; Silk *et al*., 2024) are attributable to the 1-month versus 4-month delay between the second and third vaccinations, and not the fractionation of the RH5.1 antigen dose (also noting in these trials the dose of adjuvant was not fractionated). We have previously speculated as to potential mechanisms driving improved B cell and antibody responses with delayed booster dosing (lower concentrations of circulating antibody at time of final booster and more robust germinal centre responses) and these hypotheses are under investigation in other active and upcoming RH5 vaccine clinical trials (Nielsen *et al*., 2023; Barrett, Pipini, *et al*., 2024; Bundi *et al*., 2025). This finding of comparable responses between the two regimens has important practical implications. The 10D regimen provides a substantial dose-sparing benefit and is therefore more cost-effective to produce and deliver at scale. Furthermore, it will be simpler to execute and less prone to dose-related error during a large-scale deployment.

The main limitations of this study are the sample size and demographics of the cohort. With respect to the number of participants, the relatively small numbers per group result in a lack of statistical power for between-group comparisons, as emphasised in the Statistical Analysis Plan for this study. Likewise, while we include comparisons to immunogenicity data following administration of RH5.1/AS01_B_ in the same DFx regimen in a previous Phase Ia trial in UK adults for context, we avoid over-interpretation of these findings given the small samples sizes in both trials. Future comparisons of adjuvants with larger sample sizes may be more informative. Also, although the BIO-002 study participants were predominantly from a white ethnic background, our results are in line with findings reported from the VAC080 trial (Silk *et al*., 2024) that compared delayed and delayed fractional boosting in Tanzanian children. Neither trial suggests an advantage of using the DFx dosing regimen, over the delayed regimen, with RH5.1/Matrix-M for the induction of functional anti-RH5 immunogenicity. These results thus provide important data for guiding dose selection and regimen refinement for future malaria vaccine safety and efficacy trials incorporating adjuvanted RH5.1.

## Data Availability

The original contributions presented in the study are included in the article/supplementary material, further inquiries
can be directed to the corresponding author/s.

## 5. Author Contributions

KH was the clinical lead. AMM, ROP, and SJD were chief, principal, and laboratory lead investigators in the clinical trial. CJ and IO were the lead nurses on the study with oversight of study visits in the Sheffield NIHR Clinical Research Facility. TWR, BM, TW, LK, ECB, AD, SES performed experiments or sample processing. TWR, ECB, AD, JS, CAL, SES, and CMN analysed and/or reviewed data. RA, LDWK, CC and KS worked on vaccine provision. NGM provided statistical oversight, and REC, PL, FLN, J-SC, and KS provided project management. KH and CN wrote the manuscript. All authors contributed to the article and approved the submitted version.

## 7. Acknowledgements

We thank all the vaccinees for their participation in the BIO-002 trial. The authors are grateful for the assistance of Jenny Bryant, Hannah Davies, Jing Jin, Alison Lawrie, David Pulido, Lana Strmecki (University of Oxford); Xinxue Liu, Parvinder Aley, Andrew Roberts, Chidimma Nwankwo (Oxford Vaccine Group); Catherine Green (Clinical Biomanufacturing Facility); Jenny Reimer (Novavax); Kazutoyo Miura (NIAID, NIH); and Paul Collini as the Local Safety Monitor

This work was supported primarily by the UK Medical Research Council (MRC) [Grant ref MR/V038427/1], and in part by the National Institute for Health and Care Research (NIHR) Oxford Biomedical Research Centre, the NIHR Sheffield Biomedical Research Centre, the NIHR Sheffield Clinical Research Facility. ROP also held an NIHR Academic Clinical Lecturer award [CL-2016-04-502]. This UK-funded MRC award was carried out in the frame of the Global Health EDCTP3 Joint Undertaking. JS is supported by an MRC Predoctoral Clinical Research Training Fellowship [grant number MR/Z505134/1]. The views expressed are those of the author(s) and not necessarily those of the NIHR or the Department of Health and Social Care. Matrix-M^®^ adjuvant was supplied by Novavax AB and the GIA work was supported by the Division of Intramural Research, National Institute of Allergy and Infectious Diseases, and by an Interagency Agreement (AID-GH-T-15–00001) between the US Agency for International Development Malaria Vaccine Development Program and National Institute of Allergy and Infectious Diseases. The findings and conclusions are those of the authors and do not necessarily represent the official position of US Agency for International Development. SJD is a Jenner Investigator.

