## Supplementary material for "Comparable safety and humoral immunogenicity of delayed versus delayed fractional boosting with blood-stage malaria RH5.1/Matrix-M vaccine"

### Supplementary Material for Holliday K *et al.*

#### Methods

#### Study Design

The BIO-002 trial was a single-blind, randomised trial conducted according to the current revision of the Declaration of Helsinki 2013 and in full conformity with the ICH guidelines for Good Clinical Practice (GCP). It was approved by combined review of Medicines & Healthcare products Regulatory Agency (MHRA), London-Chelsea Research Ethics Committee (REC) and Health Research Authority (HRA). (IRAS Project ID: 1005754, REC reference: 23/LO/0058). The BIO-002 trial was registered on ClinicalTrials.gov (NCT06141057) and ISRCTN95289709. The study was conducted at the NIHR Clinical Research Facility (CRF) in Sheffield Teaching Hospitals in collaboration with the University of Sheffield (Sheffield, UK) and University of Oxford (Oxford, UK).

We report here the safety, reactogenicity and immunogenicity of vaccination of RH5.1 with Matrix-M® adjuvant in delayed and delayed fractional third dose regimens up to 547 days after the first study vaccination (see **Table S1**).

#### Participants

Approved recruitment materials were sent out via University of Sheffield (UoS) volunteer staff and student mailing lists and Sheffield Teaching Hospitals (STH) NHS Trust staff newsletter. The Trust also shared the recruitment material via their social media platforms. Posters were displayed around UoS and STH premises. QR codes were provided on posters for ease. All volunteers were directed to a website with the Participant Information Sheet (PIS) and pre-screening questionnaire. Once completed, this could be accessed by study staff to confirm their interest and arrange an initial screening visit.

##### *Screening visit*

Volunteers were seen in the NIHR Clinical Research Facility (CRF), O floor, Royal Hallamshire Hospital, Sheffield. Informed consent was taken by the investigator in the clinic. The trial was explained in detail to the participants, and all had received the PIS with sufficient time to review in advance. Potential participants were consented in line with Good Clinical Practice (GCP) guidance. Any questions were answered by the investigator at the time of consent. Participants had to meet all the inclusion criteria and none of the exclusion criteria to be enrolled into the trial.

##### *Inclusion Criteria:*

- Healthy adults aged 18 to 50 years.
- Able and willing (in the Investigator's opinion) to comply with all study requirements.
- Willing to allow the Investigators to discuss the volunteer's medical history with their GP.
- Agreement to refrain from blood donation for the duration of the study.
- Able and willing to provide written informed consent to participate in the trial.
- Participants of childbearing potential only: must practice continuous effective contraception for the duration of the study. Acceptable forms of contraception include:

    - established use of oral, injected or implanted hormonal contraception
    - an intrauterine device or intrauterine system
    - male sterilisation (if the vasectomised partner is the sole partner for the participant)
    - true abstinence when in line with the preferred and usual lifestyle of the participant.
    - If a participant had experienced menarche previously and was now surgically sterile or post-menopausal, then they were deemed not of childbearing potential.

*Exclusion Criteria:*

- History of clinical malaria (any species) or previous participation in any malaria (vaccine) trial or controlled human malaria infection (CHMI) study.
- Travel to a clearly malaria endemic locality during the study period or within the preceding six months.
- Use of immunoglobulins or blood products (e.g. blood transfusion) in the last three months.
- Receipt of any vaccine in the 30 days preceding enrolment, or planned receipt of any other vaccine within 30 days following each study vaccination, with the exception of COVID-19 vaccines, which should not be received between 14 days before to 7 days after any study vaccination.
- Receipt of an investigational product in the 30 days preceding enrolment, or planned receipt during the study period.
- Concurrent involvement in another clinical trial involving an investigational product or planned involvement during the study period.
- Prior receipt of an investigational vaccine likely to impact on interpretation of the trial data, as assessed by the Investigator.
- Any confirmed or suspected immunosuppressive or immunodeficient state, including HIV infection; asplenia; recurrent, severe infections and chronic (more than 14 days) immunosuppressant medication within the past 6 months (inhaled and topical steroids were allowed).
- History of allergic disease or reactions likely to be exacerbated by any component of the vaccine.
- Any history of anaphylaxis.
- Pregnancy, lactation or intention to become pregnant during the study.
- Body mass index of <18.5 or >35.
- History of cancer (except basal cell carcinoma of the skin and cervical carcinoma in situ).
- History of serious psychiatric condition that may affect participation in the study.
- Any other serious chronic illness requiring hospital specialist supervision.
- Suspected or known current alcohol misuse as defined by an alcohol intake of greater than 25 standard UK units every week.
- Suspected or known injecting drug use in the 5 years preceding enrolment.
- Hepatitis B surface antigen (HBsAg) detected in serum.
- Seropositive for hepatitis C virus (antibodies to HCV) at screening (unless volunteer has taken part in a prior hepatitis C vaccine study with confirmed negative HCV antibodies prior to participation in that study, and negative HCV ribonucleic acid (RNA) PCR at screening for this study)

- Volunteers unable to be closely followed for social, geographic or psychological reasons.
- Any clinically significant abnormal finding on biochemistry or haematology blood tests, urinalysis or clinical examination. Blood test abnormalities will be assessed as per the laboratory adverse event (**Table S2**). In the event of abnormal test results deemed to be clinically significant, confirmatory repeat tests were requested.
- Any other significant disease, disorder, or finding which may significantly increase the risk to the volunteer because of participation in the study, affect the ability of the volunteer to participate in the study or impair interpretation of the study data.
- Inability of the study team to contact the volunteer's GP to confirm medical history and safety to participate.

#### Safety Oversight

Safety was assessed by the frequency, incidence and nature of AEs and SAEs arising during the study. The safety profile was assessed on an ongoing basis by the Investigators during the safety reporting window for each participant (from provision of consent to their last study visit). The PI/CI and relevant Investigators (as per the trial delegation log) also reviewed safety issues and SAEs as they arose.

##### *Definitions*

###### Adverse Event (AE)

An AE is any untoward medical occurrence in a volunteer, which may occur during or after administration of an Investigational Medicinal Product (IMP) and does not necessarily have a causal relationship with the intervention. An AE can therefore be any unfavourable and unintended sign (including an abnormal laboratory finding – see **Table S2**), symptom or disease temporally associated with the study intervention, whether or not considered related to the study intervention.

Each participant-reported AE was graded by the participant according to the table for grading severity of adverse events (see **Table S3**). Severity gradings were reviewed and discussed with the participants at the clinic visits. AEs were standardised and evaluated using MedDRA coding.

###### Adverse Reaction (AR)

An AR is any untoward or unintended response to an IMP. This means that a causal relationship between the IMP and an AE is at least a reasonable possibility, i.e. the relationship cannot be ruled out. All cases judged by the reporting medical Investigator as having a reasonable suspected causal relationship to an IMP (i.e. possibly, probably or definitely related to an IMP) qualified as ARs.

###### Adverse Event of Special Interest (AESI)

Adverse events identified as being of relevance to the IMP. These would have been reported as an SAE, if meeting SAE criteria (e.g. hospitalisation).

Because it has been hypothesised that immunisations with or without adjuvant may be associated with autoimmunity, regulatory authorities have requested that Novavax instruct investigators to be especially vigilant regarding the potential immune-mediated medical conditions (PIMMCs) listed in **Table S4**. Note that this regulatory request is not specific to Novavax's Matrix-M® adjuvant and there is no current evidence to suggest that the study vaccines in this protocol are, or are not, associated with these illnesses. The list is not intended to be exhaustive, nor does it exclude the possibility that other diagnoses may be an AESI.

##### Serious Adverse Event (SAE)

An SAE is an AE that results in any of the following outcomes, whether or not considered related to the study intervention:

- Death.
- Life-threatening event (i.e. the volunteer was, in the view of the Investigator, at immediate risk of death from the event that occurred). This does not include an AE that, if it occurred in a more severe form, might have caused death.
- Persistent or significant disability or incapacity (i.e. substantial disruption of one's ability to carry out normal life functions).
- Hospitalisation or prolongation of hospitalisation, regardless of length of stay, even if it is a precautionary measure for continued observation. Hospitalisation (including inpatient or outpatient hospitalisation for an elective procedure) for a pre-existing condition that has not worsened unexpectedly does not constitute an SAE.
- An important medical event (that may not cause death, be life-threatening, or require hospitalisation) that may, based upon appropriate medical judgment, jeopardise the volunteer and/or require medical or surgical intervention to prevent one of the outcomes listed above. Examples of such medical events include allergic reaction requiring intensive treatment in an emergency room or clinic, blood dyscrasias, or convulsions that do not result in inpatient hospitalisation.
- Congenital anomaly or birth defect.

##### Serious Adverse Reaction (SAR)

An AE (expected or unexpected) that is both serious and, in the opinion of the reporting Investigator or Sponsors, believed to be possibly, probably or definitely due to an IMP or any other study treatments, based on the information provided.

##### Suspected Unexpected Serious Adverse Reaction (SUSAR)

All SARs at least possibly related to RH5.1 or Matrix-M® would be considered unexpected and reported as SUSARs. Safety information relating to RH5.1 administered with adjuvants was available in the RH5.1 Investigator Brochure. There were no expected SARs for this vaccine.

##### Foreseeable Adverse Reactions

The foreseeable ARs following vaccination with RH5.1 and Matrix-M® adjuvant are: at injection site: pain, erythema, warmth, swelling, pruritus; systemically: myalgia, arthralgia, headache, fatigue, fever, feverishness, malaise and nausea. These AEs were listed as 'solicited AEs' providing they occurred within 7 days of the day of vaccination. 'Unsolicited AEs' are AEs other than the foreseeable ARs occurring within the first 7 days, or any AEs occurring after the first 7 days after vaccination.

###### *Causality Assessment*

For every unsolicited AE, an assessment of the relationship of the event to the administration of the vaccine was undertaken by CI-delegated clinician at the coordinating site (Sheffield). An intervention-related AE refers to an AE for which there is a possible, probable or definite relationship to administration of a vaccine (**Table S5**). An interpretation of the causal relationship of the intervention to the AE in question was made, based on the type of event; the temporal relationship to vaccine administration; and the known biology of the vaccine.

Causality assessment took place during planned safety reviews, interim analyses and at the final safety analysis.

*Stopping/Holding Rules*

As this was a Phase I trial safety holding rules were developed. None of these rules were triggered throughout the course of the trial. The group holding rules were:

- **Solicited local AEs:**
    - If more than two doses of vaccine were followed by a grade 3 solicited local AE beginning within 48 hours of vaccination and persisting at grade 3 for > 72 hrs.
- **Solicited systemic AEs:**
    - If more than two doses of vaccine were followed by a grade 3 solicited systemic AE beginning within 48 hours of vaccination and persisting at grade 3 for > 48 hrs.
- **Unsolicited AEs:**
    - If 2 or more volunteers developed a grade 3 unsolicited AE (including the same laboratory AE) that was considered possibly, probably or definitely related to vaccination.
    - However, if a study participant had a Grade 3 unsolicited (or laboratory) AE considered 'possibly' related to vaccination which persisted at Grade 3 for < 48 hours that, in the opinion of the Investigator, is of non-clinical significance and where a different cause is judged as likely, the event would not be counted as part of the group holding AEs.
- **An AESI or SAE considered possibly, probably or definitely related to vaccination occurred.**

In addition to group holding rules, stopping rules for individual volunteers applied (i.e., indications to withdraw individuals from further vaccinations).

- **Local reactions:** Injection site ulceration, abscess or necrosis.
- **Laboratory AEs:**
    - The volunteer developed a grade 3 laboratory AE considered possibly, probably or definitely related within 168 hours (7 days) after vaccination and persisting continuously at grade 3 for >72hrs.
- **Solicited systemic AEs:**
    - The volunteer developed a grade 3 systemic solicited AE considered possibly, probably or definitely related within 48 hours after vaccination and persisting continuously at grade 3 for >72hrs.
- **Unsolicited AEs:**
    - The volunteer had a grade 3 AE, considered possibly, probably or definitely related to vaccination, persisting continuously at grade 3 for >72hrs.
    - The volunteer had an acute allergic reaction or anaphylactic shock following the administration of the vaccine investigational product.
- **An AESI or SAE considered possibly, probably or definitely related to vaccination occurs**

In addition to these pre-defined criteria, the study could have been put on hold upon advice of the local safety monitor, CI, study sponsor, regulatory authority or REC, for any single event or combination of events which, in their professional opinion, jeopardised the safety of the volunteers or the reliability of the data.

#### Safety analysis

Following each vaccination participants visited the CRF for follow up visits on days 1, 2, 7, 14 after the first vaccination. On day 28 for the second vaccination and days 29, 35, 42, 56 for follow up. On day 182 for third vaccination visit and days 183,184, 189, 196, 210, 238, 266, 294 and 547 for follow up. Solicited AEs were collected daily for 7 days with e-diary cards. Volunteers were asked to record any AEs daily, for 28 days (unsolicited AEs). E-diaries were reviewed with volunteers at each post-vaccination clinic visit. SAEs were reported throughout the study. Heart rate, blood pressure and temperature were measured at all visits. Bloods samples for safety were taken at all visits except Day 14, 42, 56 and 196 up to Day 210. Blood samples for exploratory immunology analyses were taken at every visit except screening.

#### Primary Objective

To assess the safety of RH5.1 soluble protein with Matrix-M® in healthy adult volunteers using two different dosing regimens.

#### Secondary Objectives

1. To assess the humoral immunogenicity of RH5.1 soluble protein with Matrix-M® when administered to healthy volunteers at different doses.
2. To compare the anti-RH5 serum IgG functional immunogenicity between the groups receiving soluble RH5.1 protein at different doses in a delayed versus delayed fractional third dose regimens – assessment of serum anti-RH5 IgG quantity, functional quality and longevity.
3. To compare differences in the innate immune responses following the first and third vaccinations, and correlate these with adverse event data and adaptive immune responses.

#### Outcome Measures

##### *Primary Safety Outcome Measures*

The specific endpoints for safety and reactogenicity were actively and passively collected data on adverse events. The following parameters were assessed:

- Occurrence of solicited local reactogenicity signs and symptoms for 7 days following each vaccination
- Occurrence of solicited systemic reactogenicity signs and symptoms for 7 days following each vaccination
- Occurrence of unsolicited adverse events for 28 days following the vaccination
- Change from baseline for safety laboratory measures for 28 days following vaccination
- Occurrence of serious adverse events during the whole study duration

Solicited and unsolicited AE data were collected at each clinic visit. They were collected from e-diary cards, clinical review, clinical examination (including observations) and laboratory results. Volunteers were followed for approximately 12 - 18 months following initial trial vaccination and approximately 6 - 12 months following the third dose.

##### *Secondary Immunological Outcome Measures*

RH5-specific immunogenicity was assessed by a variety of immunological assays, with comparison before and after vaccination. The following measures were assessed:

- Serum ELISA response:
  - Quantitative antigen-specific IgG antibody levels (µg/mL readout) over time – analysis of peak responses and longevity;
  - Antigen-specific antibody subclass/isotype analysis;
  - Antigen-specific antibody avidity analysis;
- *In vitro* GIA against 3D7 clone *P. falciparum* parasites using purified total IgG and a single-cycle pLDH readout assay
- Purified IgG ELISA versus GIA titration "Quality Analysis":

#### Vaccines

RH5.1 is a full-length protein of the *Plasmodium falciparum* antigen reticulocyte-binding protein homologue 5 (PfRH5); an essential protein in the merozoite invasion of red blood cells. The design, production and pre-clinical testing has previously been reported in detail. It was produced to current Good Manufacturing Practice (cGMP) standards using a *Drosophila melanogaster* Schneider 2 (S2) cell line by the Clinical Biomanufacturing Facility (CBF), University of Oxford. Fill-and-finish was completed at Symbiosis, Scotland and labelled and released by PCI Pharma services to Sheffield Teaching Hospitals NHS Foundation Trust pharmacy.

The protein was supplied as a liquid formulation in a Tris buffer at a concentration of 0.2 ± 0.025 mg/mL in a volume of 500 µL. The drug product was a clear, colourless, frozen liquid essentially free of particles, supplied in 2 mL sterile glass vials.

RH5.1 was stored in a locked, temperature-controlled freezer at -80°C (nominal). All movements and administration of the vaccine were documented in an accountability log.

Matrix-M® is a licensed saponin-based adjuvant system manufactured by Novavax AB (Uppsala, Sweden). PCI Pharma services certified and labelled before supplying to Sheffield Teaching Hospitals NHS Foundation Trust pharmacy.

Matrix-M® is formulated at a concentration of 0.375 mg/mL in PBS with a fill volume of 0.75 mL.

#### Peripheral Blood Mononuclear Cell (PBMC), Plasma and Serum preparation

All bloods taken for exploratory immunology purposes were labelled with participant ID, date, and time of sample. In addition, a paper sample log was completed for each participant that recorded the time of transit to lab, volume of samples and time of processing. These were taken to University of Sheffield (UoS) laboratories and processed for storage and shipping by the laboratory technical team in UoS. Bloods were processed as per the local SOP.

Briefly, for serum samples this involved 1 h standing at room temperature (RT), then spinning at 1300 x g for 5 min. Serum was then collected with transfer pipette, aliquoted and stored at -80°C.

For PBMC isolation, all processing was done within 6 h of blood sampling. 20 – 30 mL of heparinised blood was poured into one prepared Leucosep tube. For blood volumes greater than 30 mL, this was split between two or more prepared Leucosep tubes each with 15 mL Lymphoprep below the porous filter disc. This was then centrifuged at 1000 x g for 13 min at RT (Brake setting 0, Accel setting 2). Samples were then collected of the plasma fraction as required into labelled 2 mL cryovials and/or 15 mL falcons and stored at -80°C.

The remaining liquid containing PBMC and excess plasma from each tube was then poured or pipetted into a new, labelled 50 mL falcon, topped up with R0 media (RPMI 1640, L-glutamine and Pen/Strep) and spun at 700 x g for 5 min at RT (Brake setting 9, Accel setting 9). One wash tube was used per Leucosep tube. The supernatant was poured off and the pellet resuspended in 5 mL RBC Lysis solution. Cells were then rested for a maximum of 5 min and topped up to 30 mL with R0, spun at 700 x g for 5 min at RT (Brake setting 9, Accel setting 9). If RBC contamination was extensive, this step was repeated.

The supernatant was then poured off and the pellet resuspended in 10 mL R10 (RPMI 1640, L-glutamine, Pen/Strep and 10% foetal bovine serum (FBS)) for counting.
Cells were then counted using the DeNovix CellDrop Counter. After this, cells were resuspended initially in cold FBS at 0.5 mL per aliquot then topped up to 1 mL with 20% DMSO in FBS and stored in 1 mL aliquots of ~6 million cells/cryovial. Initially these were put in a CoolCell in the -80°C freezer and then later transferred to the Biorepository for long term storage in liquid nitrogen. Some of these samples were later shipped to the University of Oxford for analysis.

### Results

#### Solicited Adverse Events

For further clarity, a table detailing the maximum grade of each symptom after each vaccine with frequency and proportions is provided in **Table S6**.

#### Unsolicited Adverse Events

In addition to that reported in the main paper, the rest of the unsolicited AEs are listed in **Table S7**. Most of these were mild or moderate with only four events graded as severe. One participant in the DFx group reported migraine after the second vaccine that lasted less than a day, with a past medical history of the same. An episode of fainting occurred the morning after the second vaccine for one participant in the 10D group. The same participant also reported calf pain that lasted 48 hours after the third vaccine. A further participant reported back pain that lasted 1 day after the second vaccine in the 10D group.

#### Laboratory Adverse Events

Lab AEs were graded as per **Table S2** and the safety bloods were performed by the clinical laboratories at Royal Hallamshire Hospital, Sheffield Teaching Hospitals NHS Trust.

### Tables

***Table S1 Illustration of vaccination doses and timepoints***

| Group | Number of participants | Day 0 | Day 28 | Day 182 |
|---|---|---|---|---|
| Delayed third dose "10D" | 12 | RH5.1 10µg Matrix-M® 50µg | RH5.1 10µg Matrix-M® 50µg | RH5.1 10µg Matrix-M® 50µg |
| Delayed fractional third dose "DFx" | 12 | RH5.1 50µg Matrix-M® 50µg | RH5.1 50µg Matrix-M® 50µg | RH5.1 10µg Matrix-M® 50µg |

***Table S2 Safety bloods laboratory AE grading definitions***

| | | | Grade 0 | NCS | Grade 1 | Grade 2 | Grade 3 |
|---|---|---|---|---|---|---|---|
| Haemaglobin | **Male** | g/l | 166 - 131 | 129-126 | 125 - 115 | 114 - 100 | <100 |
| | **Female** | | 147 - 110 | - | 109 - 100 | 99 - 90 | <90 |
| | **Hb change from baseline** | | | | Oct-15 | 16 - 20 | 21 - 50 |
| White Blood Cells **Elevated** | **Male** **Female** | $x10^9/l$ | 3.5 - 9.5 | 9.6 - 9.9 | 10.0 - 15.0 | 15.1-20.0 | >20 |
| White Blood Cells **Low** | **Male** **Female** | $x10^9/l$ | 3.5 - 9.5 | 3.3 - 3.4 | 2.5 - 3.2 | 1.5 - 2.4 | <1.5 |
| Platelets | **Male** **Female** | $x10^9/l$ | 150 - 400 | 141 - 149 | 110 - 140 | 95-109 | <95 |
| Neutrophils | | $x10^9/l$ | 1.7 - 6.5 | - | 1.00-1.69 | 0.50-0.99 | <0.50 |
| Lymphocytes | | $x10^9/l$ | 1.0 - 3.0 | - | 0.75 - 0.99 | 0.50-0.74 | <0.50 |
| Eosinophils | | $x10^9/l$ | 0.04 - 0.5 | 0.51-0.59 | 0.60 - 1.50 | 1.51 - 5.00 | >5.00 |
| | | | **Grade 0** | **NCS** | **Grade 1** | **Grade 2** | **Grade 3** |
| Sodium | **Elevated** | mmol/l | 133-146 | - | 147 | 148-149 | >149 |
| | **Low** | | | | 130-132 | 128-129 | <128 |
| Potassium | **Elevated** | mmol/l | 3.5-5.3 | - | 5.4-5.5 | 5.6-5.7 | >5.7 |
| | **Low** | | | | 3.2-3.3 | 3.0-3.1 | <3.0 |
| Urea | | µmol/l | 2.5-7.8 | 7.9-8.1 | 8.2-8.9 | 9.0-11.0 | >11.0 |
| Creatinine | Female | µmol/l | 44 - 80 | 81 - 87 | 1.1-1.5*ULN | >1.5-3.0*ULN | >3.0*ULN |
| | | | | | *88 - 120* | *121 - 240* | *>240* |
| Creatinine | Male | µmol/l | 62 - 106 | 107 - 116 | 1.1-1.5*ULN | >1.5-3.0*ULN | >3.0*ULN |
| | | | | | *117 - 159* | *160 - 318* | *>318* |
| Bilirubin | **Normal LFT** | µmol/l | 0- 21 | 22 - 24 | 1.2-1.5*ULN | >1.5-2.0*ULN | >2.0*ULN |
| | | | | | *25-32* | *33-42* | *>42* |
| Bilirubin | **Abnormal LFT** | µmol/l | 0 - 21 | 22 | 1.1-1.25*ULN | >1.25-1.5*ULN | >1.5-1.75*ULN |
| | | | | | *23-26* | *27-32* | *>32* |
| ALT | Female | IU/l | 0 - 33 | 34 - 35 | 1.1-2.5*ULN | >2.5-5.0*ULN | >5.0*ULN |
| | | | | | *36 - 83* | *84 - 165* | *>165* |
| | Male | IU/l | 0 - 41 | 42 - 44 | 1.1-2.5*ULN | >2.5-5.0*ULN | >5.0*ULN |
| | | | | | *45 - 103* | *104 - 205* | *>205* |
| Alk Phosphatase | | IU/l | 30 - 130 | 131 - 142 | 1.1-2.0*ULN | >2.0-3.0*ULN | >3.0*ULN |
| | | | | | *143-260* | *261-390* | *>390* |
| Albumin | | g/l | 35 - 50 | 34 - 32 | 31 - 28 | 27 - 25 | <25 |
| | | | **Grade 0** | **NCS** | **Grade 1** | **Grade 2** | **Grade 3** |
| Prothombin Time (PT) | | s | 9.5 - 11.5 | 11.6 - 11.9 | 1.05 - 1.10*ULN | 1.11-1.20*ULN | 1.21*ULN |
| | | | | | *12.0 - 12.7* | *12.8 - 13.8* | *>13.8* |
| Activated Partial Thromboplastin Time (APTT) | | s | 20.0 - 27.1 | 33-34 | 1.10-1.20*ULN | >1.20-1.4*ULN | >1.40*ULN |
| | | | | | *29.8 - 32.5* | *32.6 - 37.9* | *>37.9* |
| Fibrinogen | | g/l | 2.0 - 4.0 | 1.5 - 1.99 | 1.25-1.49 | 1.0-1.24 | <1.0 ** |

***Table S3 Severity grading criteria for AEs***

| Grade | Severity |
|---|---|
| Grade 0 | None |
| Grade 1 | Mild: Transient or mild discomfort (< 48 h); no medical intervention/therapy required |
| Grade 2 | Moderate: Mild to moderate limitation in activity – some assistance may be needed; no or minimal medical intervention/therapy required |
| Grade 3 | Severe: Marked limitation in activity, some assistance usually required; may require medical intervention/therapy |

***Table S4 Listings of Adverse Events of Special Interest (AESIs)/ Potential Immune-Mediated Medical Conditions (PIMMCs)***

| Categories | Diagnoses (as MedDRA Preferred Terms) |
|---|---|
| Neuroinflammatory Disorders: | Acute disseminated encephalomyelitis (including site-specific variants: e.g., non-infectious encephalitis, encephalomyelitis, myelitis, cranial nerve disorders including paralyses/paresis (e.g., Bell's palsy), generalized convulsion, Guillain-Barre syndrome (including Miller Fisher syndrome and other variants), immune-mediated peripheral neuropathies and plexopathies (including chronic inflammatory demyelinating polyneuropathy, multifocal motor neuropathy and polyneuropathies associated with monoclonal gammopathy), myasthenia gravis, multiple sclerosis, narcolepsy, optic neuritis, transverse myelitis, uveitis. |
| Musculoskeletal and Connective Tissue Disorders: | Antisynthetase syndrome, dermatomyositis, juvenile idiopathic arthritis (including Still's disease), connective tissue disorder, polymyalgia rheumatic, polymyositis, psoriatic arthropathy, polychondritis, rheumatoid arthritis, scleroderma (including diffuse systemic form and CREST syndrome), spondyloarthritis (including ankylosing spondylitis, reactive arthritis [Reiter's Syndrome] and undifferentiated spondyloarthritis), systemic lupus erythematosus, systemic sclerosis, Sjogren's syndrome. |
| Vasculitides: | Large vessels vasculitis (including giant cell arteritis such as Takayasu's arteritis and temporal arteritis), medium sized and/or small vessels vasculitis (including polyarteritis nodosa, Kawasaki's disease, microscopic polyangiitis, Wegener's granulomatosis, Churg–Strauss syndrome [allergic granulomatous angiitis], Buerger's disease [thromboangiitis obliterans], necrotizing vasculitis and ANCA-positive vasculitis [type unspecified], Henoch-Schonlein purpura, Behcet's syndrome). |
| Gastrointestinal Disorders: | Crohn's disease, celiac disease, ulcerative colitis, ulcerative proctitis. |
| Hepatic Disorders: | Autoimmune hepatitis, autoimmune cholangitis, cholangitis sclerosing primary biliary cirrhosis. |

| | |
|---|---|
| Renal Disorders: | Autoimmune glomerulonephritis (including IgA nephropathy, glomerulonephritis rapidly progressive, membranous glomerulonephritis, membranoproliferative glomerulonephritis, and mesangioproliferative glomerulonephritis). |
| Cardiac Disorders: | Autoimmune myocarditis/cardiomyopathy. |
| Skin Disorders: | Alopecia areata, psoriasis, vitiligo, Raynaud's phenomenon, erythema nodosum, autoimmune bullous skin diseases (including pemphigus, pemphigoid and dermatitis herpetiformis), cutaneous lupus erythematosus, morphoea, lichen planus, Stevens-Johnson syndrome. |
| Hematologic Disorders: | Autoimmune haemolytic anaemia, immune thrombocytopenia, antiphospholipid syndrome, thrombocytopenia. |
| Metabolic Disorders: | Autoimmune thyroiditis, Basedow's disease, Hashimoto thyroiditis, diabetes mellitus type 1, Addison's disease. |
| Other Disorders: | Goodpasture syndrome, idiopathic pulmonary fibrosis, pernicious anaemia, sarcoidosis. |

Abbreviations: ANCA = anti-neutrophil cytoplasmic antibody; IgA = immunoglobulin A; MedDRA = Medical Dictionary for Regulatory Activities.

***Table S5: Guidelines for assessing the relationship of vaccine administration to an AE.***

| | |
|---|---|
| No Relationship | No temporal relationship to study product ***and***<br>Alternate aetiology (clinical state, environmental or other interventions); ***and***<br>Does not follow known pattern of response to study product |
| Unlikely | Unlikely temporal relationship to study product ***and***<br>Alternate aetiology likely (clinical state, environmental or other interventions) ***and***<br>Does not follow known typical or plausible pattern of response to study product. |
| Possible | Reasonable temporal relationship to study product; ***or***<br>Event not readily produced by clinical state, environmental or other interventions; ***or***<br>Similar pattern of response to that seen with other vaccines |
| Probable | Reasonable temporal relationship to study product; ***and***<br>Event not readily produced by clinical state, environment, or other interventions ***or***<br>Known pattern of response seen with other vaccines |

| | |
|---|---|
| Definite | Reasonable temporal relationship to study product; ***and***<br>Event not readily produced by clinical state, environment, or other interventions; ***and***<br>Known pattern of response seen with other vaccines |

***Table S6 Frequency and proportion of maximum severity of each symptom reported, faceted by group and by dose***

| Symptom | None | | Mild | | Moderate | | Severe | |
|---|---|---|---|---|---|---|---|---|
| | 10D | DFx | 10D | DFx | 10D | DFx | 10D | DFx |
| **Dose 1** | N (%) | N (%) | N (%) | N (%) | N (%) | N (%) | N (%) | N (%) |
| Fatigue | 6 (50%) | 6 (50%) | 5 (41.7%) | 5 (41.7%) | 1 (8.3%) | 1 (8.3%) | 0 (0%) | 0 (0%) |
| Fever | 11 (91.7%) | 12 (100%) | 1 (8.3%) | 0 (0%) | 0 (0%) | 0 (0%) | 0 (0%) | 0 (0%) |
| Feverish | 11 (91.7%) | 12 (100%) | 0 (0%) | 0 (0%) | 1 (8.3%) | 0 (0%) | 0 (0%) | 0 (0%) |
| Headache | 7 (58.3%) | 9 (75%) | 5 (41.7%) | 3 (25%) | 0 (0%) | 0 (0%) | 0 (0%) | 0 (0%) |
| Itch | 12 (100%) | 12 (100%) | 0 (0%) | 0 (0%) | 0 (0%) | 0 (0%) | 0 (0%) | 0 (0%) |
| Joint Pain | 11 (91.7%) | 10 (83.3%) | 1 (8.3%) | 2 (16.7%) | 0 (0%) | 0 (0%) | 0 (0%) | 0 (0%) |
| Muscle Pain | 6 (50%) | 7 (58.3%) | 6 (50%) | 3 (25%) | 0 (0%) | 2 (16.7%) | 0 (0%) | 0 (0%) |
| Nausea | 8 (66.7%) | 10 (83.3%) | 3 (25%) | 2 (16.7%) | 1 (8.3%) | 0 (0%) | 0 (0%) | 0 (0%) |
| Pain | 4 (33.3%) | 6 (50%) | 8 (66.7%) | 6 (50%) | 0 (0%) | 0 (0%) | 0 (0%) | 0 (0%) |
| Redness | 12 (100%) | 12 (100%) | 0 (0%) | 0 (0%) | 0 (0%) | 0 (0%) | 0 (0%) | 0 (0%) |
| Swelling | 12 (100%) | 12 (100%) | 0 (0%) | 0 (0%) | 0 (0%) | 0 (0%) | 0 (0%) | 0 (0%) |
| Unwell | 8 (66.7%) | 10 (83.3%) | 3 (25%) | 2 (16.7%) | 1 (8.3%) | 0 (0%) | 0 (0%) | 0 (0%) |
| Warmth | 8 (66.7%) | 10 (83.3%) | 3 (25%) | 2 (16.7%) | 1 (8.3%) | 0 (0%) | 0 (0%) | 0 (0%) |
| **Dose 2** | | | | | | | | |
| Fatigue | 6 (50%) | 6 (50%) | 2 (16.7%) | 1 (8.3%) | 4 (33.3%) | 5 (41.7%) | 0 (0%) | 0 (0%) |
| Fever | 10 (83.3%) | 11 (91.7%) | 2 (16.7%) | 1 (8.3%) | 0 (0%) | 0 (0%) | 0 (0%) | 0 (0%) |
| Feverish | 5 (41.7%) | 5 (41.7%) | 6 (50%) | 2 (16.7%) | 1 (8.3%) | 5 (41.7%) | 0 (0%) | 0 (0%) |
| Headache | 3 (25%) | 4 (33.3%) | 5 (41.7%) | 4 (33.3%) | 4 (33.3%) | 4 (33.3%) | 0 (0%) | 0 (0%) |
| Itch | 10 (83.3%) | 9 (75%) | 2 (16.7%) | 3 (25%) | 0 (0%) | 0 (0%) | 0 (0%) | 0 (0%) |
| Joint Pain | 5 (41.7%) | 6 (50%) | 3 (25%) | 6 (50%) | 4 (33.3%) | 0 (0%) | 0 (0%) | 0 (0%) |
| Muscle Pain | 3 (25%) | 2 (16.7%) | 5 (41.7%) | 5 (41.7%) | 3 (25%) | 5 (41.7%) | 1 (8.3%) | 0 (0%) |

| | | | | | | | | |
|---|---|---|---|---|---|---|---|---|
| Nausea | 7 (58.3%) | 8 (66.7%) | 3 (25%) | 2 (16.7%) | 0 (0%) | 2 (16.7%) | 2 (16.7%) | 0 (0%) |
| Pain | 1 (8.3%) | 1 (8.3%) | 9 (75%) | 7 (58.3%) | 2 (16.7%) | 4 (33.3%) | 0 (0%) | 0 (0%) |
| Redness | 9 (75%) | 12 (100%) | 3 (25%) | 0 (0%) | 0 (0%) | 0 (0%) | 0 (0%) | 0 (0%) |
| Swelling | 11 (91.7%) | 11 (91.7%) | 1 (8.3%) | 1 (8.3%) | 0 (0%) | 0 (0%) | 0 (0%) | 0 (0%) |
| Unwell | 5 (41.7% | 7 (58.3%) | 3 (25%) | 1 (8.3%) | 3 (25%) | 4 (33.3%) | 1 (8.3%) | 0 (0%) |
| Warmth | 8 (66.7%) | 8 (66.7%) | 4 (33.3%) | 3 (25%) | 0 (0%) | 1 (8.3%) | 0 (0%) | 0 (0%) |
| **Dose 3** | | | | | | | | |
| Fatigue | 1 (9.1%) | 4 (33.3%) | 3 (27.3%) | 6 (50%) | 6 (54.5%) | 1 (8.3%) | 1 (9.1%) | 1 (8.3%) |
| Fever | 9 (81.8%) | 12 (100%) | 2 (18.2%) | 0 (0%) | 0 (0%) | 0 (0%) | 0 (0%) | 0 (0%) |
| Feverish | 4 (36.4%) | 9 (75%) | 5 (45.5%) | 3 (25%) | 2 (18.2%) | 0 (0%) | 0 (0%) | 0 (0%) |
| Headache | 3 (27.3%) | 6 (50%) | 5 (45.5%) | 4 (33.3%) | 3 (27.3%) | 1 (8.3%) | 0 (0%) | 1 (8.3%) |
| Itch | 8 (72.7%) | 7 (58.3%) | 3 (27.3%) | 4 (33.3%) | 0 (0%) | 1 (8.3%) | 0 (0%) | 0 (0%) |
| Joint Pain | 4 (36.4%) | 7 (58.3%) | 3 (27.3%) | 4 (33.3%) | 4 (36.4%) | 1 (8.3%) | 0 (0%) | 0 (0%) |
| Muscle Pain | 3 (27.3%) | 2 (16.7%) | 4 (36.4%) | 8 (66.7%) | 4 (36.4%) | 1 (8.3%) | 0 (0%) | 1 (8.3%) |
| Nausea | 5 (45.5%) | 9 (75%) | 4 (36.4%) | 2 (16.7%) | 2 (18.2%) | 1 (8.3%) | 0 (0%) | 0 (0%) |
| Pain | 1 (9.1%) | 4 (33.3%) | 6 (54.5%) | 6 (50%) | 4 (36.4%) | 2 (16.7%) | 0 (0%) | 0 (0%) |
| Redness | 8 (72.7%) | 9 (75%) | 3 (27.3%) | 3 (25%) | 0 (0%) | 0 (0%) | 0 (0%) | 0 (0%) |
| Swelling | 11 (100%) | 8 (66.7%) | 0 (0%) | 4 (33.3%) | 0 (0%) | 0 (0%) | 0 (0%) | 0 (0%) |
| Unwell | 6 (54.5%) | 9 (75%) | 1 (9.1%) | 1 (8.3%) | 4 (36.4%) | 2 (16.7%) | 0 (0%) | 0 (0%) |
| Warmth | 8 (72.7%) | 9 (75%) | 3 (27.3%) | 2 (16.7%) | 0 (0%) | 1 (8.3%) | 0 (0%) | 0 (0%) |

366

**Table S7. Summary of all unsolicited adverse events reported within 28 days of each vaccine deemed possibly, probably or definitely related to the vaccine.**

| Unsolicited adverse events | Group 1 | | | | Group 2 | | | |
|---|---|---|---|---|---|---|---|---|
| | V1 | V2 | V3 | **Max Grade** | V1 | V2 | V3 | **Max Grade** |
| Cough | 0 | 1 | 0 | Mild | 0 | 0 | 0 | NA |
| Decreased appetite | 0 | 0 | 1 | Moderate | 0 | 0 | 0 | NA |
| Dizziness | 1 | 0 | 0 | Mild | 0 | 0 | 0 | NA |
| Dysgeusia | 1 | 0 | 0 | Mild | 0 | 0 | 0 | NA |
| Fainting | 0 | 1 | 0 | Severe | 0 | 0 | 0 | NA |
| Feverish (post-D7) | 1 | 0 | 0 | Moderate | 0 | 0 | 0 | NA |
| Gastrointestinal disturbance | 0 | 1 | 2 | Moderate | 0 | 0 | 1 | Moderate |
| Generalised itching | 0 | 0 | 0 | NA | 1 | 0 | 0 | Mild |
| Headache | 1 | 1 | 0 | Moderate | 2 | 1 | 0 | Severe |
| Hot feeling in feet | 0 | 0 | 1 | Moderate | 0 | 0 | 0 | NA |
| Injection site bruising/swelli | 1 | 1 | 1 | Moderate | 3 | 0 | 0 | Moderate |
| Injection site itching/warmth | 2 | 2 | 0 | Mild | 1 | 1 | 0 | Mild |
| Lymphadenopathy | 0 | 0 | 1 | MIld | 1 | 2 | 0 | Moderate |
| Numbness in toes | 1 | 0 | 0 | Mild | 0 | 0 | 0 | NA |
| Paraesthesia upper limb* | 1 | 1 | 1 | Mild | 0 | 0 | 0 | NA |
| Rash trunk | 1 | 0 | 0 | Mild | 0 | 0 | 0 | NA |
| Rhinitis | 0 | 1 | 0 | Moderate | 1 | 0 | 0 | Mild |
| Restless Sleep | 0 | 0 | 1 | Mild | 0 | 0 | 0 | NA |
| Sweating easily | 0 | 0 | 1 | Mild | 0 | 0 | 0 | NA |
| Feet swelling | 0 | 0 | 1 | Mild | 0 | 0 | 0 | NA |

* Two are the same participant after different doses

### Figures

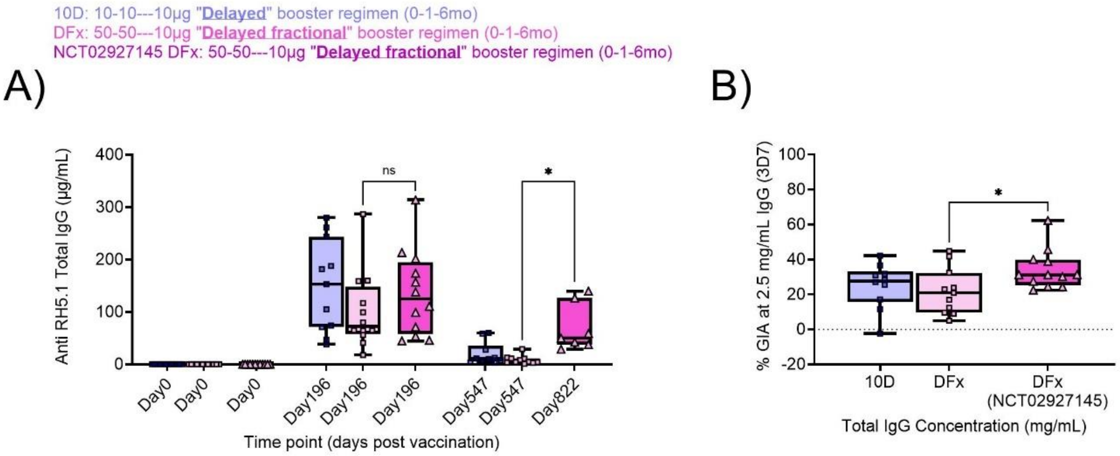


**Supplementary Figure 1. Magnitude and quality of RH5.1-specific IgG response to delayed fractional boosting with RH5.1/Matrix-M® in comparison to delayed fractional boosting with RH5.1/AS01$_B$.** Anti-RH5.1 serum IgG kinetics from pre-vaccination baseline (Day 0), peak (Day 196), and late time point (Day 547 or Day 822) were measured by ELISA (**A**). A Kruskal Wallis test was performed to compare serum IgG concentrations between BIO-002 study Group 2 (G2) DFx (delayed fractional RH5.1/Matrix-M®) and G3 DFx (NCT02927145 delayed fractional RH5.1/AS01$_B$) at Day 0, Day 196, and Day 547/ 822. *In vitro* GIA was analysed with total IgG purified from peak post-vaccination (Day 196; V3+14) serum. GIA activity at 2.5 mg/mL was compared between G2 DFx and NCT02927145 G3 DFx participants by Mann Whitney (**B**). In all graphs, Group 1 (10D) data are shown in blue, Group 2 (DFx) in pink, and NCT02927145 Group 3 (DFx) in dark pink. Each point represents a single sample. Bars and error bars represent median and interquartile ranges, respectively. p values are annotated on graphs; $p < 0.05$ was considered significant. ns = non-significant. * $p < 0.05$. Full details of the NCT02927145 clinical trial can be found in the primary trial publication (Minassian *et al.*, 2021).
